# The mortality due to COVID-19 in different nations is associated with the demographic character of nations and the prevalence of autoimmunity

**DOI:** 10.1101/2020.07.31.20165696

**Authors:** Bithika Chatterjee, Rajeeva Laxman Karandikar, Shekhar C. Mande

**Affiliations:** National Centre for Cell Science, NCCS Complex, Ganeshkhind, Pune- 411007; Chennai Mathematical Institute, H1 SIPCOT IT Park, Siruseri- 603103; Council of Scientific and Industrial Research, 2, Rafi Marg, Anusandhan Bhavan, New Delhi- 110001

**Keywords:** SARS-CoV-2, Risk factors, Mortality, Epidemiology, Autoimmunity, Hygiene

## Abstract

In the first few months of its deadly spread across the world, Covid-19 mortality has exhibited a wide range of variability across different nations. In order to explain this phenomenon empirically, we have taken into consideration all publicly available data for 106 countries on parameters like demography, prevalence of communicable and non-communicable diseases, BCG vaccination status, sanitation parameters etc. We ran multivariate linear regression models to find that the incidence of communicable diseases correlated negatively while demography, improved hygiene and higher incidence of autoimmune disorders correlated positively with Covid-19 mortality and were among the most plausible factors to explain Covid-19 mortality as compared to the GDP of the nations.

## Introduction

Higher GDP and improved Human Development Index (HDI) has led to improved sanitation and consequently reduction in the load of communicable disease burden in many countries^1^. Interestingly, disease burden of non-communicable diseases now occupies areas of major concern in the higher HDI countries. Thus, a distinct correlation between HDI status of a country, and the prevalence of specific diseases has emerged in recent history^2^.

About half of the world’s population lives in low and low middle income countries^3^. Typically access to healthcare facilities, hygiene and sanitation is poorer in these countries and is often believed to be the contributing factor of higher incidence of communicable diseases in these countries. Thus, it is not unexpected if infectious disease pandemics, such as that due to SARS-CoV-2, have catastrophic consequences in the low and low-middle income countries. Yet on the contrary, the disease prevalence and the Case Fatality Ratio (CFR) during the Covid-19 pandemic shows a contrasting opposite trend in the low and low-middle income countries when compared to that of the high income countries. It is fascinating to explore reasons that would explain higher prevalence and deaths due to Covid-19 among richer nations.

An interesting relationship between severity of Covid-19 outcome and several non-communicable disorders such as diabetes, hypertension, cardiovascular disorders has been noted^4–6^. However, the non-communicable disease burden of a country and its apparent relation to CFR due to Covid-19 has not been explored in detail yet. As a large population with these disorders lives in the high HDI countries, co-morbidities with diabetes, hypertension, cardiovascular disorders and respiratory disorders might have emerged as important determinants of CFR due to Covid-19 in these countries. Similarly, people above the age of 65 are also believed to be at a greater risk^7^, with the percentage of such people being significantly more in the higher HDI countries^8^. Thus, co-morbidities with non-communicable diseases and the fraction of people living above the age of 65, being skewed towards the high-income countries, offers possible explanations to the perplexing observation of CFR dichotomy among nations.

Among the many parameters that lead to non-communicable diseases, autoimmunity occupies an important place. Interestingly, a correlation between autoimmune disorders and HDI has been proposed^9^. Since, one of the primary manifestations of Covid-19 has been a severe autoimmune reaction in the later phase of the disease^10^, susceptibility to autoimmunity in SARS-CoV-2 infection is a possibility to consider. One of the reasons of rising prevalence in autoimmune disorders in the western countries has been proposed to be that related to the “hygiene hypothesis” ^11^. The hypothesis postulates that exposure to pathogens early in life protects people from allergic diseases later in their lives. Moreover, improvement in hygiene practices such as better sanitation, availability of safe drinking water, hand washing facilities etc. reduces the impact of communicable diseases. On the contrary such a reduction to the exposure to infectious agents might be related to higher prevalence of autoimmune disorders^12^. We wished to explore if epidemiological data supports correlation between autoimmune diseases’ prevalence, sanitation parameters and how much of the variation in death per million due to Covid-19 is explained by the same. We have chosen more than 25 parameters and attempted to find correlation between these and CFR, if any. These parameters include GDP, HDI, prevalence of various diseases, demographic parameters, various sanitation parameters etc. Our findings reveal that the apparent correlation between GDP of a nation and Covid-19 related deaths, or that between GDP and CFR, is a manifestation of other parameters.

## Methods

The country-wise Covid-19 deaths per million data was collected from https://ourworldindata.org/coronavirus^13^ with the original source: European Centre for Disease Prevention and Control (ECDC) on the 29^th^ June 2020. We chose deaths per million as compared to the confirmed reported cases as it gives a more reliable parameter to assess the current pandemic. The parameters on obesity (BMI>30, Age and sex standardized) was sourced from the WHO^14^ while the parameters on GDP per capita, population density, age profile, gender ratio, HDI and the urban population percentage was sourced from the World Bank organization^15^.

We also collected all possible variables that quantify the cleanliness parameters in countries. This included the percentage of access to handwashing facilities, access to basic drinking water, availability of basic sanitation, prevalence of open defecation, safe drinking and safe sanitation. Most of these variables were available through WHO and UNICEF water and sanitation surveillance projects^16^. The prevalence percentages of different autoimmune disorders such as Multiple Sclerosis, Type1 Diabetes Mellitus, Psoriasis, Rheumatoid Arthritis, Asthma, and communicable diseases such as Schistosomiasis, Onchocerciasis, Lymphatic Filariasis, Ascaris, Hook worm, Malaria, Tuberculosis, Dengue, Upper respiratory infections, Lower respiratory infections and H influenzae type B meningitis for the year 2017(Age and sex standardized) were downloaded from the http://ghdx.healthdata.org/gbd-results-tool^17^. The diseases were classified into two broad categories as communicable and non-communicable diseases.

The Mean BCG vaccination coverage in percentage was downloaded from WHO^18^. The average mean coverage was calculated for all years. The 2017 GDP gross domestic products per capita based on purchasing power parity converted to 2011 international dollars and Human Development Index 2018 was used for analysis. For details of sources of each variable see Supplementary Dataset S1.

We further filtered countries that had at least 10 deaths in total and population size of at least 100,000. Moreover, we restricted consideration of the countries to have at least 4 deaths/million. For the countries missing corresponding values for our variables, we either imputed them with values smaller than the smallest values in the other countries or dropped the countries from further analyses. For the missing values in variables related to drinking water, sanitation and wash we predicted them using a regression model as we noticed a high correlation amongst each other. For missing values of HDI, we used GDP data to impute values for HDI. After filtration and imputation of missing values we were left with 106 countries with values corresponding to more than 25 variables. For the same countries we downloaded the deaths per million up to 60 days from the onset of the first death occurring in each country on 7^th^ August 2020. Subsequently we also added the 90 days and 120 days interval data collected on 16^th^ September 2020 to account for the difference in the stage of the epidemic and the lockdown restrictions of each country. It is customary to consider log of GDP instead of GDP for analyses in many disciplines to brings it closer to normal distribution. We also considered log of deaths per million instead of deaths per million for all our calculations.

We compared individual (Pearson) correlation coefficients of different variables with the log of deaths per million (LDM) on the 29^th^ June as well as with the 60, 90 and 120 days interval LDM. The variables were compared with log (GDP) and HDI as well. This helped in assessing the important variables to be used in our linear regression model. The country-wise data on various parameters: GDP, HDI, sanitation parameters, prevalence of various communicable and non-communicable diseases were clubbed into development variables (GDP, HDI), demographics (percentages of older population, urban population and obesity among adults), Sanitation (percentage of access to handwashing facilities, basic drinking, basic sanitation, no open defecation, safe drinking and safe sanitation), communicable diseases (Schistosomiasis, Onchocerciasis, Lymphatic Filariasis, Ascaris, Hook worm, Malaria, Tuberculosis, Dengue, Upper respiratory infections, Lower respiratory infections and H influenzae type B meningitis) and non-communicable diseases (Multiple Sclerosis, Type1 Diabetes Mellitus, Psoriasis, Rheumatoid Arthritis and Asthma).

### Statistical Analysis used

To explore which of these combined variables are important, we tried multivariate linear regression model using R with several combinations of variables to obtain insights into our understanding of epidemiology of Covid-19. LDM of both scenarios was assessed for its association with our predictor/explanatory variables grouped as described above into development variables, demographic variables, sanitation, tropical diseases and autoimmune disorders. Our final objective was to select the best combination of these groups of variables that would yield the largest adjusted-R^2^ value.

## Results

Analysis was carried on reported numbers of total 10112754 cases 501562 deaths as on the 29^th^ June 2020 as well as with 60-day interval data as on the 7^th^ August 2020. Further 90 days and 120 days interval data as on 16^th^ September 2020 was also included. Data from well-known corona virus global deaths tracking sources^19–21^ show that more than 70 percent Covid-19 mortality has occurred in high income countries like Italy, Spain, UK, France and USA^22^. Our analyses also reveal a similar trend (Fig. 1a, b). To explore the dichotomy of rich and poor countries in relation to Covid-19 deaths, we considered several potential explanatory parameters. The factors included percentage of older population, population density, incidence of communicable and non-communicable diseases etc. The data shows a positive correlation between log (GDP per capita) and log (deaths per million), abbreviated as LDM (Fig. 3, Supplementary, Tables S1, Fig. S1b), which reinforces the information of richer countries being most burdened by the disease.

**Figure 1a.**
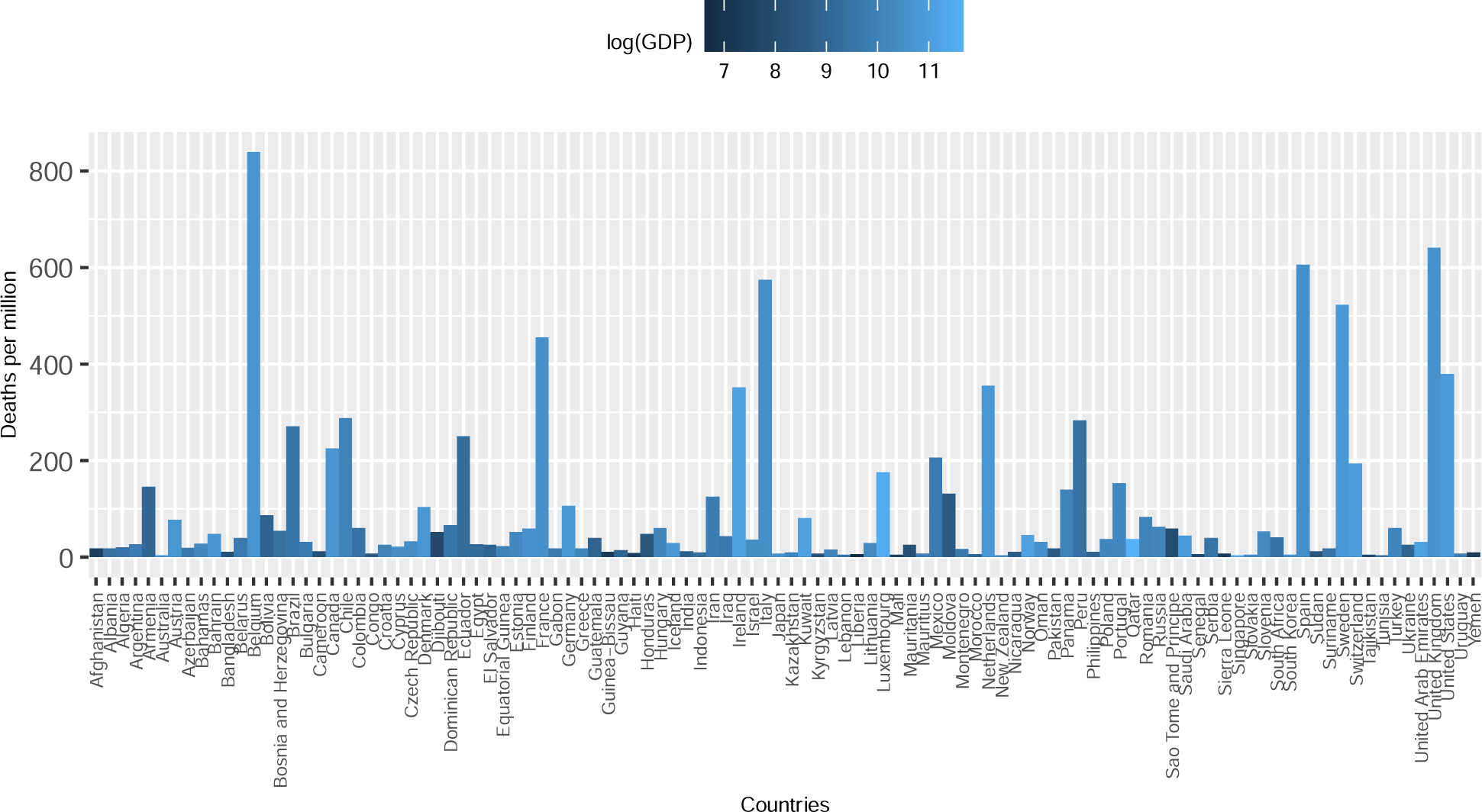
The number of Deaths per Million inhabitants in each country due to Covid-19 up to 29^th^ June 2020. Color gradient indicates the country’s log (GDP).

**Figure 1b.**
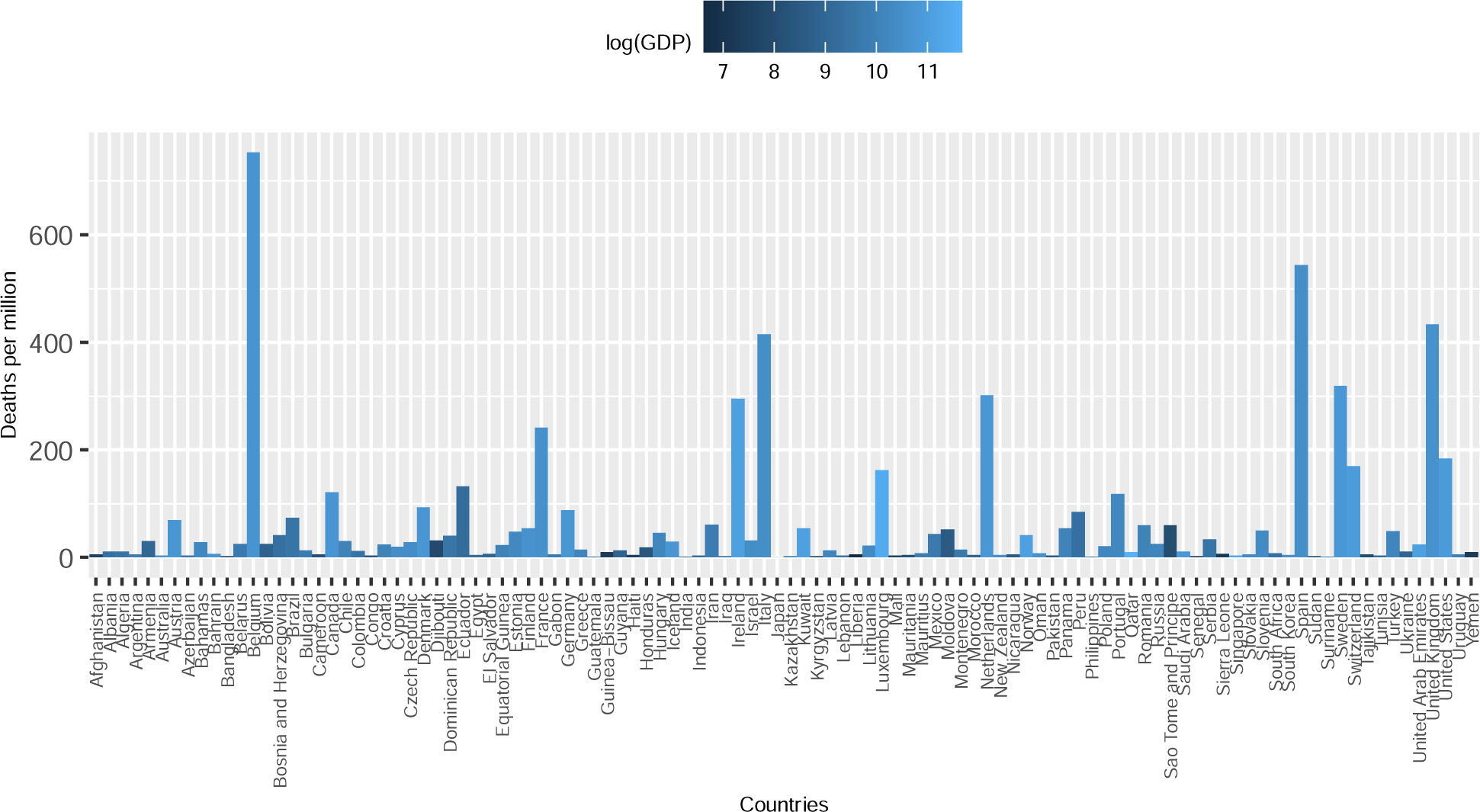
The number of Deaths per Million inhabitants in each country due to Covid-19 60 days after onset of 1^st^ death. Color gradient indicates the country’s log (GDP).

**Figure 2.**
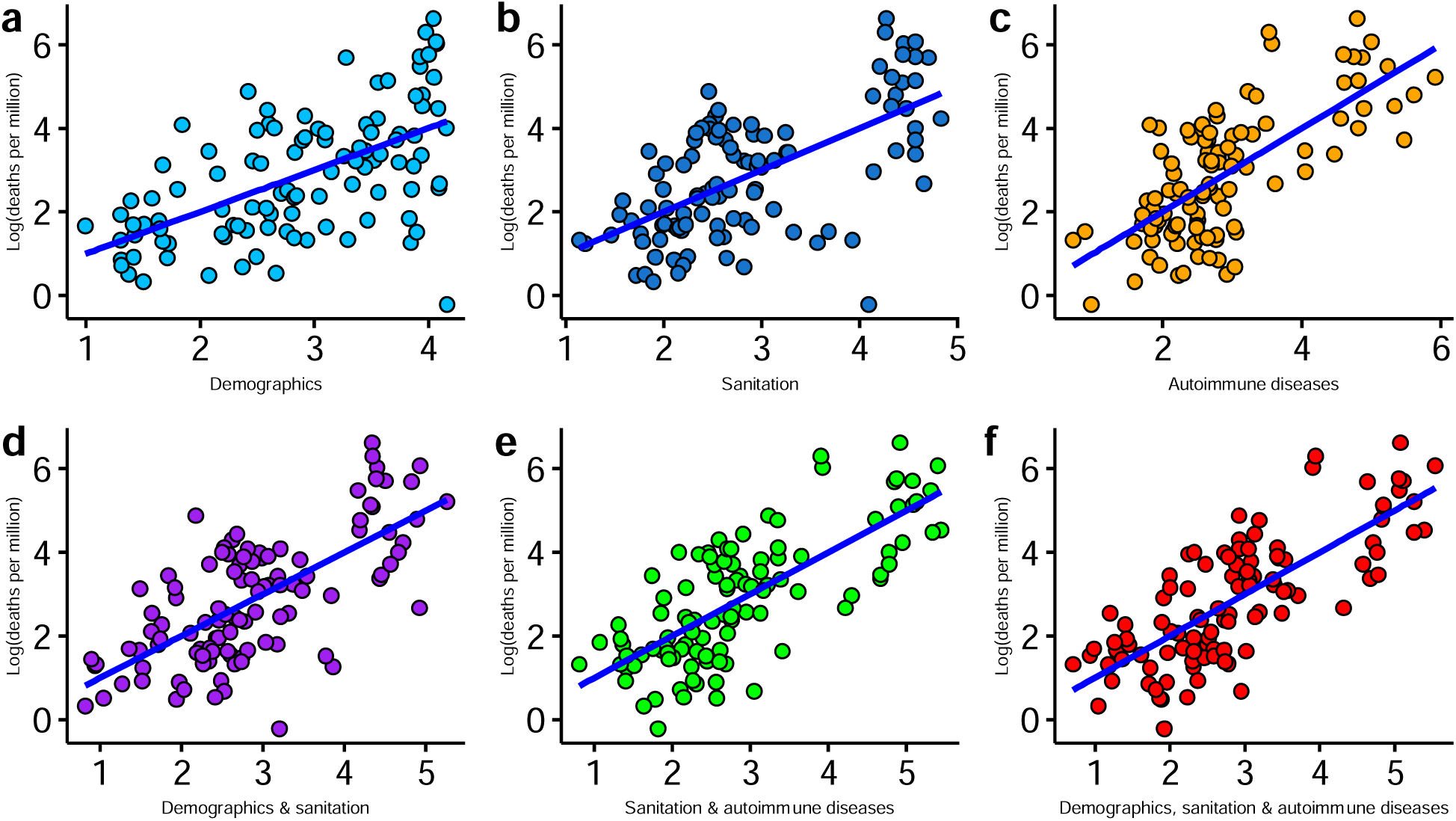
The actual values of log (deaths per million) 60 days after onset of 1^st^ death or LDM plotted against their predicted values. The explanatory variables have been grouped, as described in the text, into a) demographics, b) sanitation, c) autoimmune diseases, d) demographics & sanitation, e) sanitation & autoimmune diseases and f) demographics, sanitation and autoimmune diseases. Regression analysis were done with LDM as dependent variable and the combinations of different variables; N=106. Each dot represents a country. The R^2^ of each graph is depicted in Table 1.

**Figure 3.**
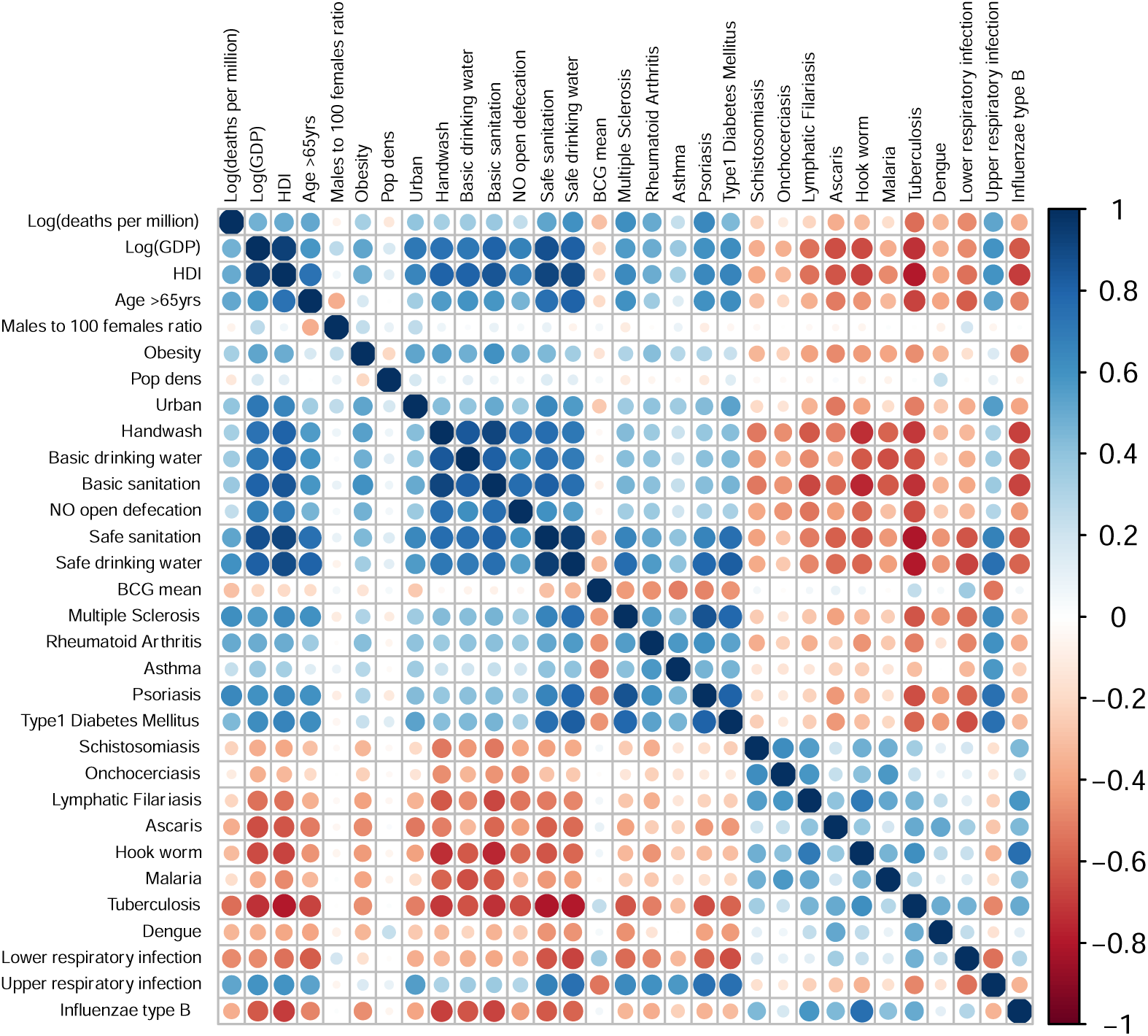
Plot showing correlation matrix of every variable with each other. The color key is depicted as −1(red) showing inverse correlation, 0 showing no correlation, and +1(blue) showing maximum positive correlation. The sizes of the circles range from small (low correlation) to larger (high correlation). Here the Log (deaths per million) is from the 60 days interval data; N=106.

**Table 1.**
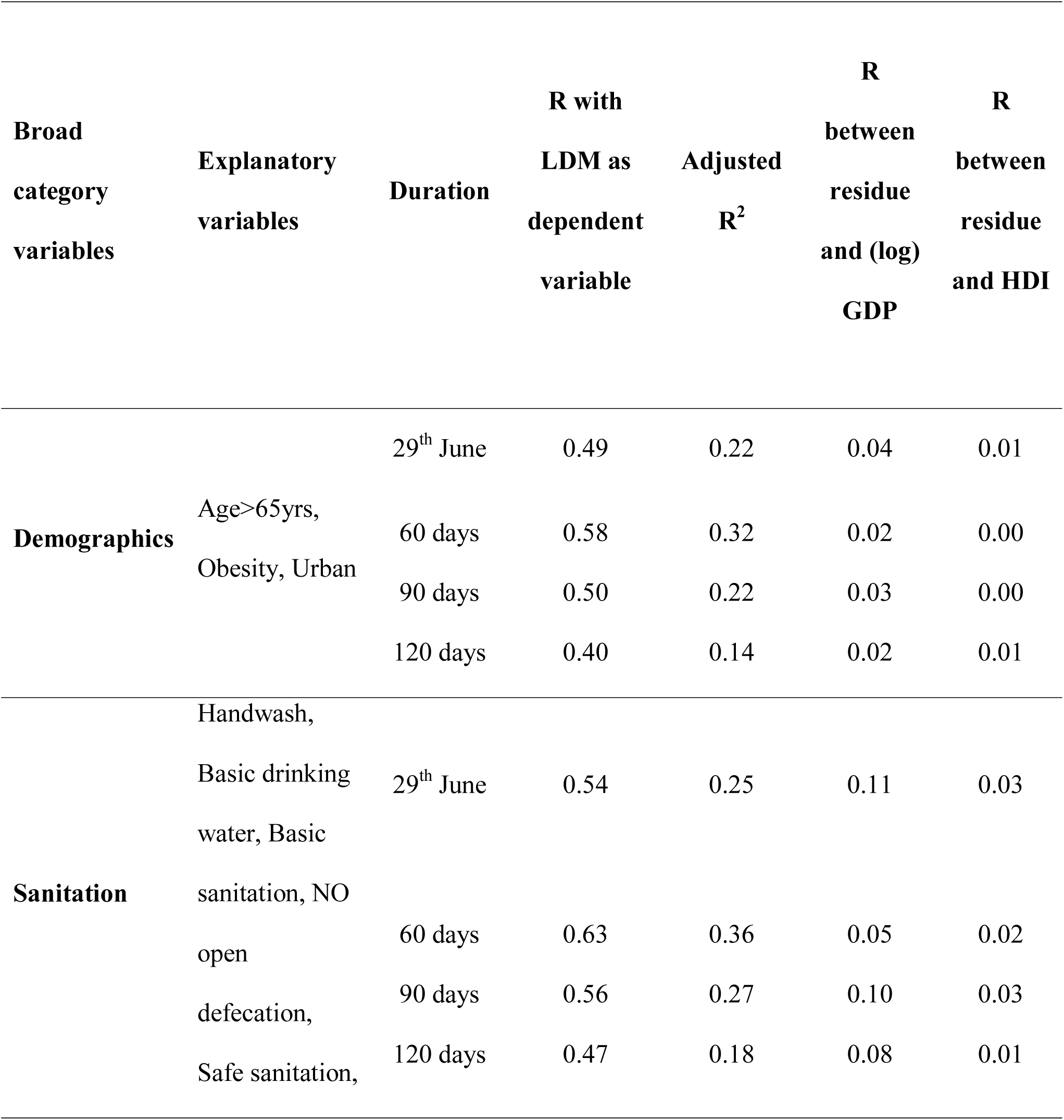
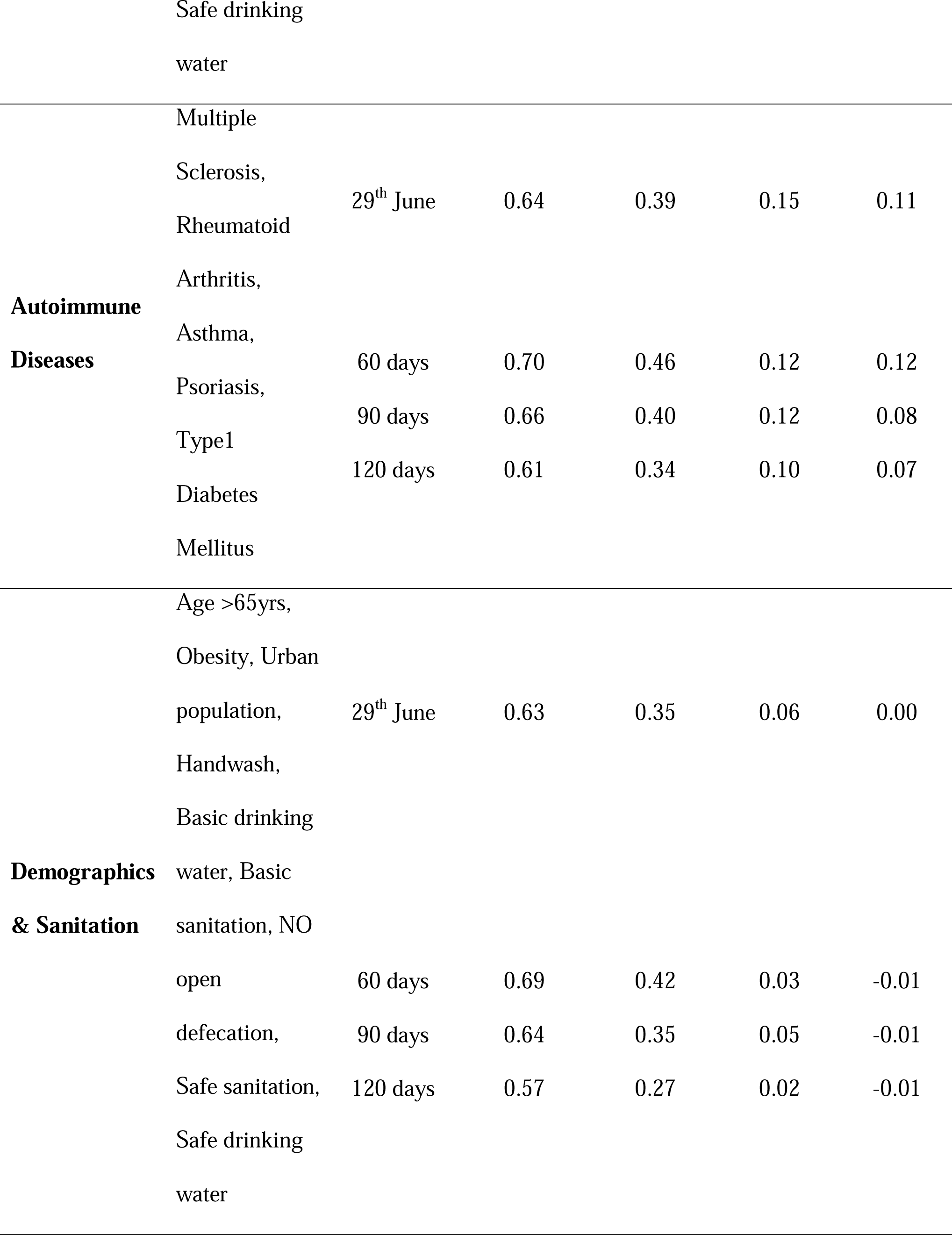
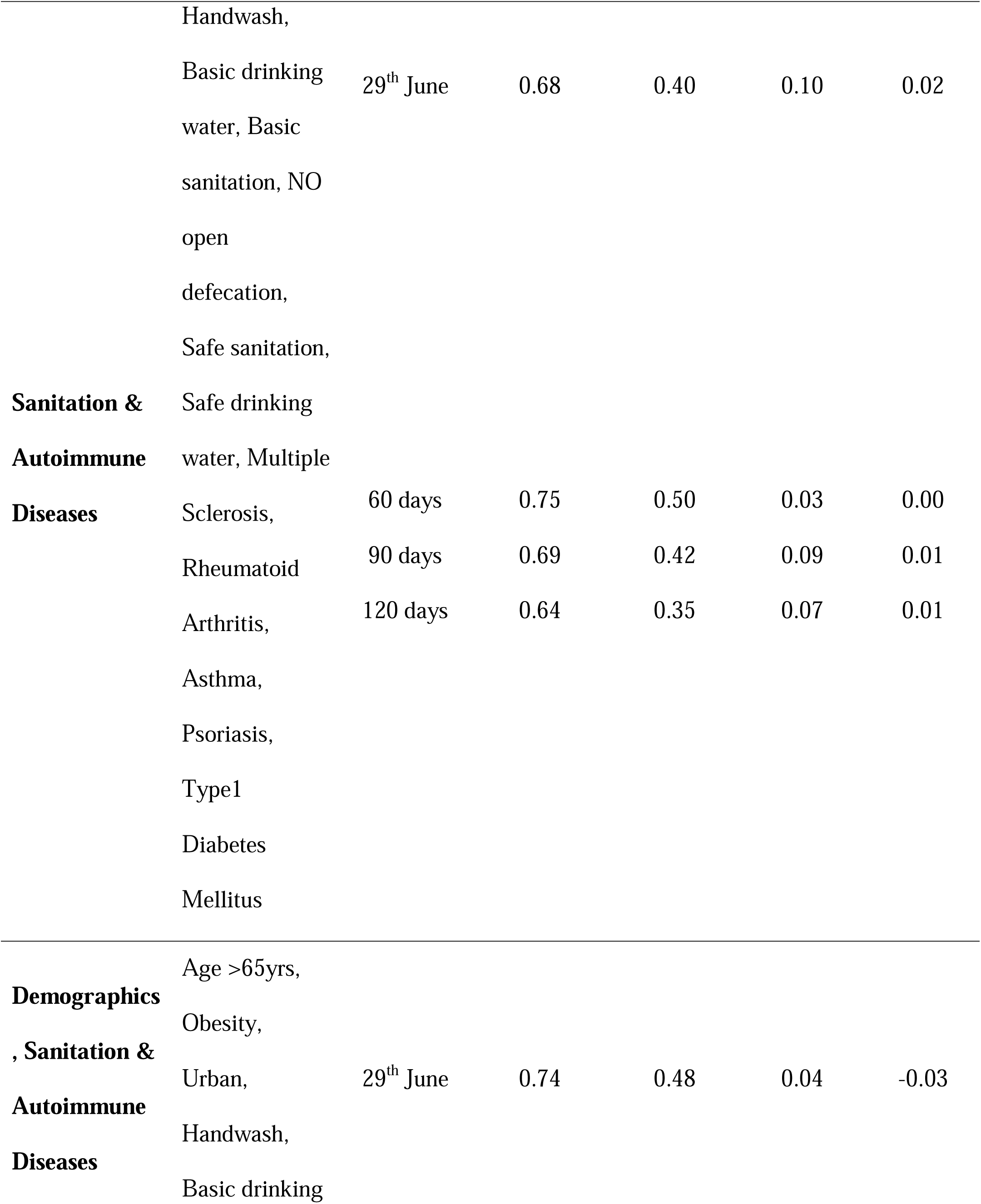
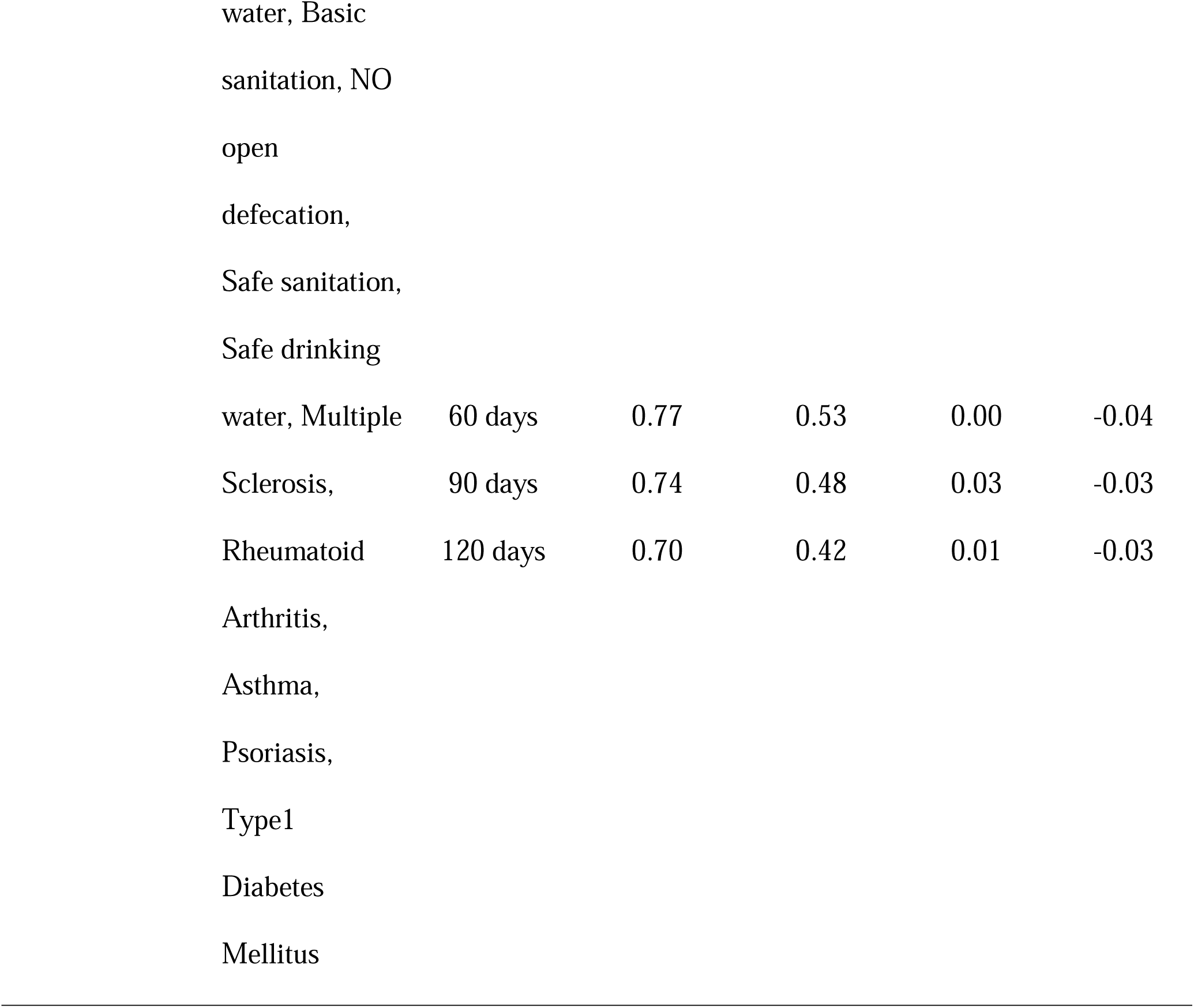
R-the correlation coefficient of combinations of variables with log (Deaths/Million) LDM for 60,90 and 120 days interval and 29^th^ June based on multiple regression analysis as wellas correlation of the residues with log (GDP) and HDI. Also showing the adjusted R^2^; N=106.

### Demographics

Among the many distinctive features of higher GDP countries, are its ageing population and a larger fraction of urbanization. We observe that while proportion of population above 65, obesity and the percentage of urban population are positively correlated with the Covid-19 deaths (Fig. 3, Supplementary, Tables S1, Fig.S1c, Fig. S1e, Fig. S1g) the gender ratio and population density have negligible negative correlation (Fig. 3, Supplementary, Tables S1, Fig.S1d, Fig. S1f). Interestingly, whereas at an individual level gender does seem to have an impact on Covid-19 related death as numerous studies have shown^23,24^, at the aggregate level the gender ratio varies little across countries. Likewise, urbanization has an impact while population density does not seem to have an impact. We observe a positive correlation between LDM and percentage of population with age over 65, which is expected from the reports showing that older population is more susceptible to the virus^7^. Thus, the overall demographic parameters yield a positive correlation with Covid-19 deaths per million (Fig. 2a).

### Safe Sanitation

Lack of sanitation and poorer hygiene practices are known to be responsible for the higher communicable disease burden in the low GDP countries^1^. It is therefore reasonable to expect that parameters describing safe sanitation and safe drinking water to be correlated negatively with the Covid-19 deaths. Surprisingly we find a contrary observation, where different sanitation parameters are correlated positively with the Covid-19 outcome (Fig. 2b). It is therefore perplexing to note positive correlation of sanitation parameters, as described in methods, with the Covid-19 CFR (Fig. 3, Supplementary, Tables S1, Fig. S1h-m).

### Communicable Diseases

Several studies have reported the protective role of parasitic/bacterial infections in bolstering human immunity, also referred to as immune training^25,26^. We observe that the prevalence of communicable diseases such as Malaria and Tuberculosis as well as parasitic diseases such as schistosomiasis, Onchocerciasis have a weak negative correlation with LDM (Fig. 3, Supplementary, Tables S1, Fig. S1t-z1). Upper respiratory diseases however show positive correlation which may be indicative of hyper active immune response in individual suffering from such disorder(Fig. 3, Supplementary, Tables S1). These intuitive observations related to communicable diseases with respect to Covid-19 deaths are suggestive of the role of “immune training” and the final disease outcome.

### BCG vaccination

Recent studies have suggested that the countries in which the majority population was immunized by BCG vaccine had lower LDM due to trained immunity^27,28^. We find negligible of correlation between BCG vaccination in different countries with LDM (Fig. 3, Supplementary, Tables S1, Fig. S1n). A possible explanation for this difference could be due to the fact that comparisons in the previous studies were made with socially similar nations while we consider a diverse group of countries. Moreover, the distribution showing two distinct clusters for BCG vaccination (Supplementary, Fig. S1n), and with no apparent correlation within the clusters also makes it difficult to draw any conclusions on the effect of BCG vaccination on CFR at the population level. It is also pertinent to note that we find significant negative correlation of mean BCG vaccination of a country and prevalence of autoimmune disorders (Fig. 3).

### Autoimmunity

One of the manifestations of better hygiene and safe sanitation practices in high GDP countries is the increased incidence in autoimmune disorders^29^. It has been postulated that better hygiene practices could lower a person’s immunity and make the person susceptible to autoimmune diseases. We therefore tested if any of the autoimmune diseases showed an association with the LDM variable. We interestingly find a strong positive correlation with Multiple Sclerosis, Type 1 Diabetes Mellitus, Rheumatoid Arthritis and Psoriasis and a weaker correlation with Asthma (Fig. 2c, Fig. 3, Supplementary, Tables S1, Fig. S1o-s).

### Regression Analysis

We now combined many related factors as described in methods such as demographic parameters, sanitation, communicable and non-communicable diseases. These combinations were arrived at after calculating correlations among all different parameters under consideration (Fig3, Supplementary, Tables S1). In order to inspect whether development indices like HDI and GDP of countries had an actual impact on our variable of LDM, we ran regression models using combinations of explanatory factors outlined above and checked the correlation between the residues, i.e. difference in actual values of the dependent variable and its predicted values from predictor variables, to GDP and HDI.

After testing various combinations of these groups, we observe that demographic variables, sanitation parameters and incidence of autoimmune disorders together explain most of the observed variation in LDM and development variables do not seem to count once the effect of these explanatory variables is accounted for (Supplementary, Tables S2). More precisely, if one uses multiple regression to predict death rates, then the residues are uncorrelated with GDP and HDI. The R value for our final best model for 60 days interval is 0.77 and the adjusted R^2^ value is 0.53(P-value<0.05). The correlation of this model’s residue with GDP and HDI came to 0.00 and −0.04 respectively (Supplementary, Tables S2). Since in this duration there was strict lockdown imposed upon some of the countries, we also did the same analysis on 90 and 120 days interval data. The R value for 90 days interval came to 0.74 and adjusted R^2^ came to 0.48. The analysis on the 120 days interval reduced R values to 0.70 and adjusted R^2^ value to 0.42. Although still significant the reduced R^2^ and R values as the number of days increases can be attributed to the lowering of deaths in countries whose first wave of infection was completed as well as better preparedness to fight the infection.

## Discussion

Various corona virus global deaths tracking sources^19–21,30^ point towards uneven distribution of Covid-19 deaths with richer countries having higher CFR. In order to understand the reasons behind this paradoxical observation we analyzed publicly available data on various parameters. In this context, it is well known that when we have multiple factors correlated with dependent variable and these factors are themselves mutually correlated, it is not possible to determine as to which of these may be the causal variables using purely statistical techniques. To address this, we could collect data from a controlled experiment or look for data on individuals rather than countrywide aggregated data. However, in this instance many variables make sense only at the aggregate level ruling out both these options. Moreover, trends observed at individual level can be opposite to ecological or population level owing to the Simpson’s paradox. In such challenging situations, it is desirable to use domain knowledge and identify the variables whose correlation with the dependent variables has a logical explanation. In the present context, older population having higher death rate can be explained biologically. Similarly, since Covid-19 spreads rapidly via person-to-person contact, population density is likely to have a big impact.

Our observation of the weak negative correlation of Covid-19 LDM with communicable diseases, and its positive correlation with incidence of autoimmune disorders in the high GDP countries, is indication of the interplay of host immunity and viral infection. As the parasite and bacterial disease burden is high in low and low-middle income countries, this can best be inferred upon by the “immune training” in the population of these countries due to chronic exposure to communicable diseases. It has been observed recently that patients susceptible to the Corona virus exhibited impaired Type 1 interferon activity^31^. Since initially the innate immunity acts upon the virus mediated by the interferon activity, individual with prior infections tend to coup with the virus better than the ones without exposure to pathogens^32^. The evidence for the innate immunity playing major role in susceptibility to the virus is available from the fact that asymptotic patients or patients with mild infection produced poor neutralizing antibodies^33^. Hence it can be speculated that viral susceptibility is established at the first line of defense known as innate immunity much before the adaptive immunity. As mentioned earlier, the observed correlation between incidence of autoimmune disorders and LDM can be explained as Covid-19 seems to lead to severe autoimmune reaction. Paradoxically, better sanitation leads to poorer “immune training” and thus could be leading to higher LDM. Thus, the observed correlation of demographics, autoimmune disorders and sanitation with LDM appears to have explanation coming from the domain. The transfer rate for COVID-19 being high, we see a lower correlation between incidence of communicable diseases and LDM due to COVID-19 as compared to autoimmune disorders.

As can be seen in (Supplementary, Tables S2), the correlation of GDP and HDI with each model’s residue is almost negligible. Both GDP and HDI seem to have a dominant role in determining the deaths per million yet both become insignificant when our linear regression models considered other notable and explainable confounding factors such as the demography, sanitation and the pre-Covid prevalence’s of autoimmune and tropical diseases. The parameters of sanitation and autoimmune diseases consistently contributed significantly to the R^2^ no matter which combination of variables were chosen. It has not escaped out attention that there could be other factors such as the country’s stage of the epidemic and lower reporting/testing in less developed countries that could also affect the mortality numbers. However, the statistical evidence given by us should give a head start to investigate the role of hyper immune reaction on Covid-19 susceptibility at individual patient’s level.

We note that statistical analyses such as this attempt to provide evidence of practices of on a macroscopic scale. Individual level variations among people, or communities, typically get masked in such analyses. Thus, although we provide a possible explanation based on sanitation practices on the CFR differences among economically stronger and weaker countries, this should not be inferred as our advocating a move towards weaker hygiene practices for handling future pandemics. Rather this analysis opens up avenues to consider “immune training” with possibilities of microbiome therapies to supplement improved hygiene and sanitation practices.

## Data Availability

dataset is available for data.

## Acknowledgements

BC gratefully acknowledges financial support by the National Centre for Cell Science, Pune. Authors declare no competing financial interests.

