## Supplementary 1 for "The mortality due to COVID-19 in different nations is associated with the demographic character of nations and the prevalence of autoimmunity"

**This Document file includes:**

Figures S1

Figures S2

Tables S1 to S2

**Other supplementary materials for this manuscript include the following:**

Dataset S1

Fig. S1.

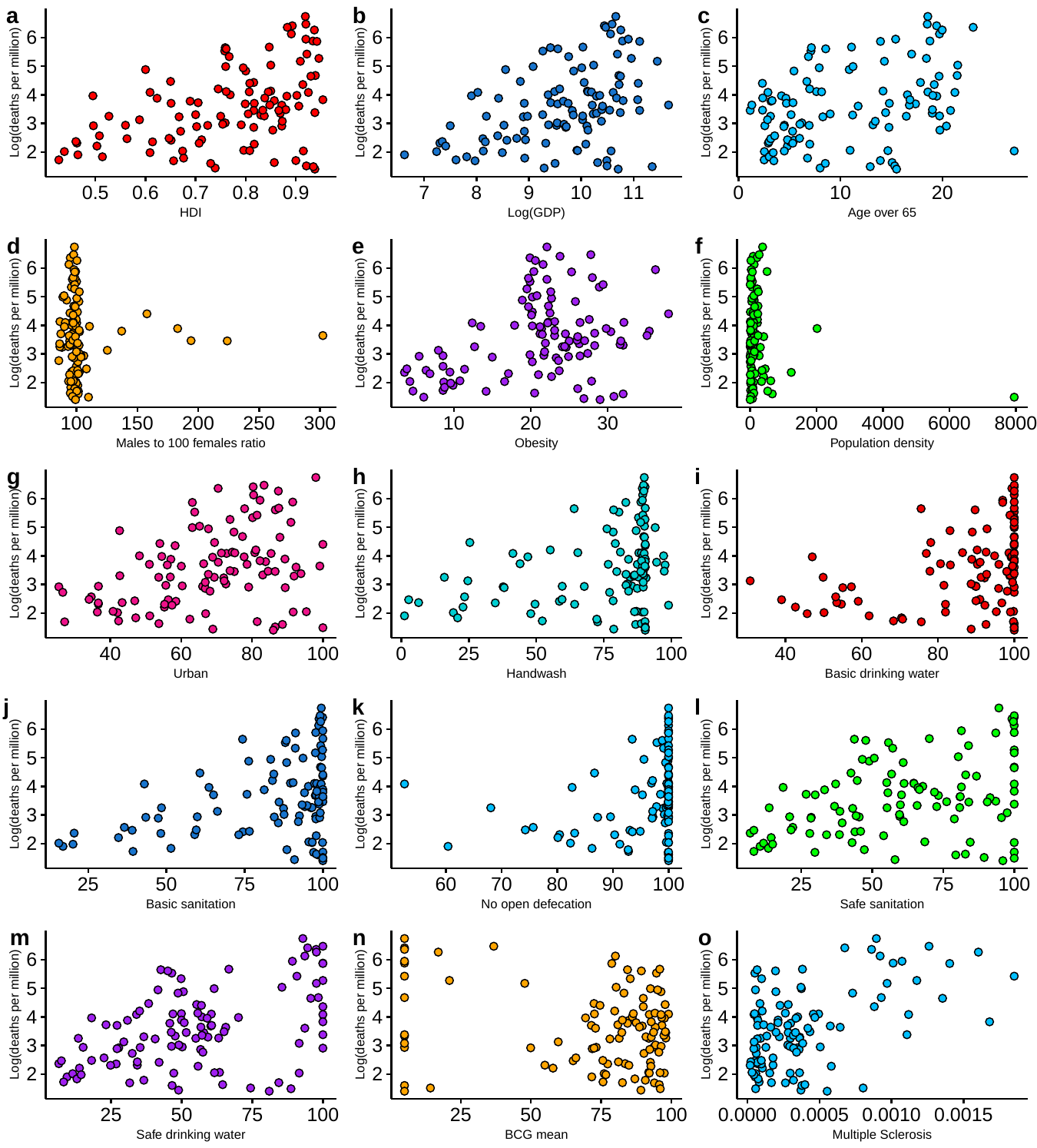

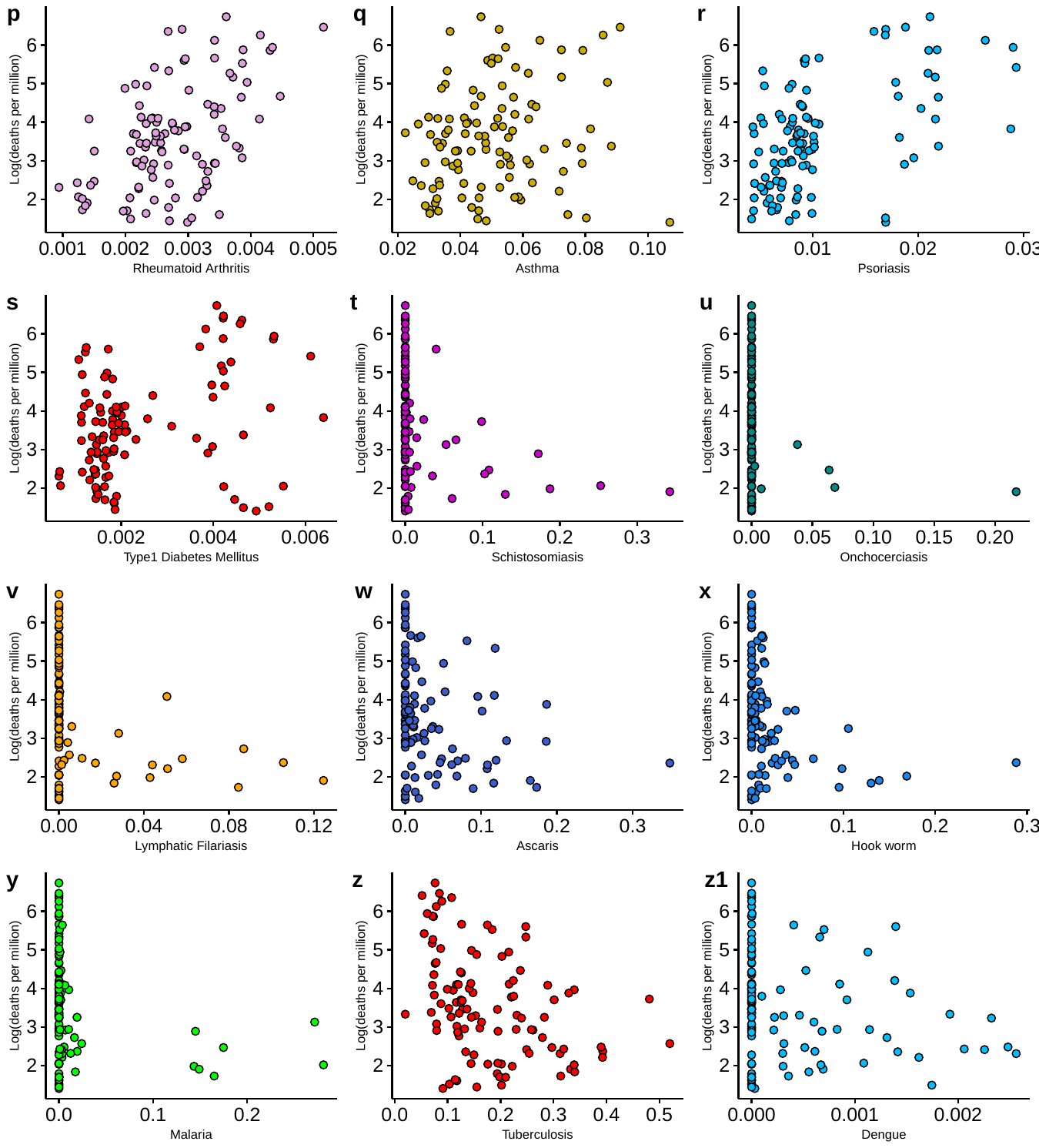

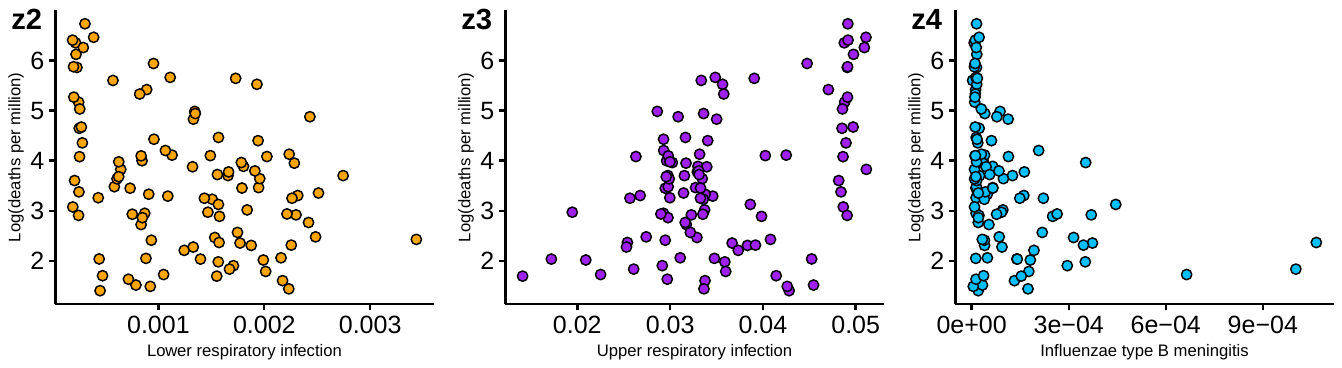

**Fig: S1** Scatter plot of each variables with log (deaths per million) as on 29^th^ June; N=106.

**Fig: S2**

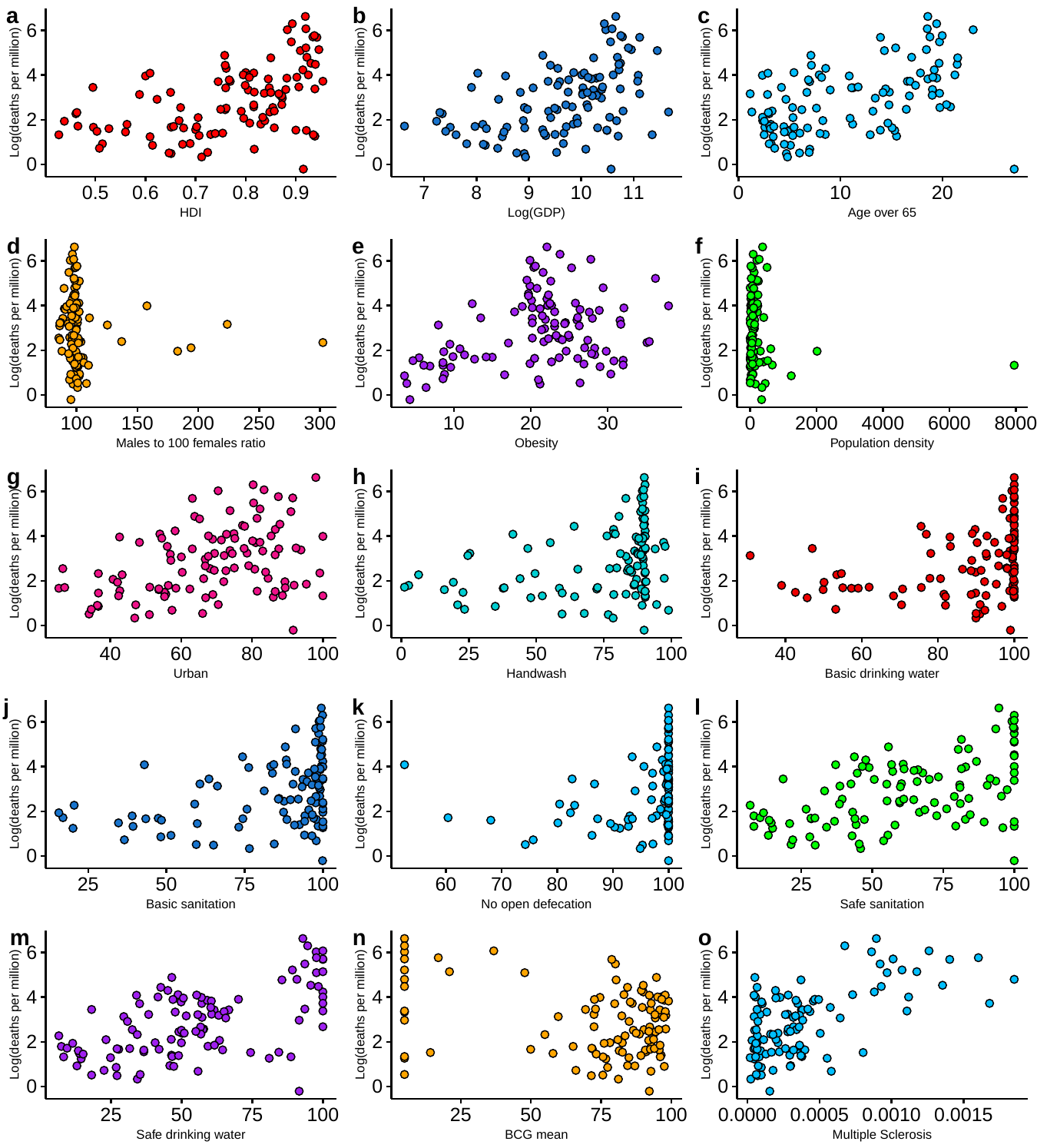

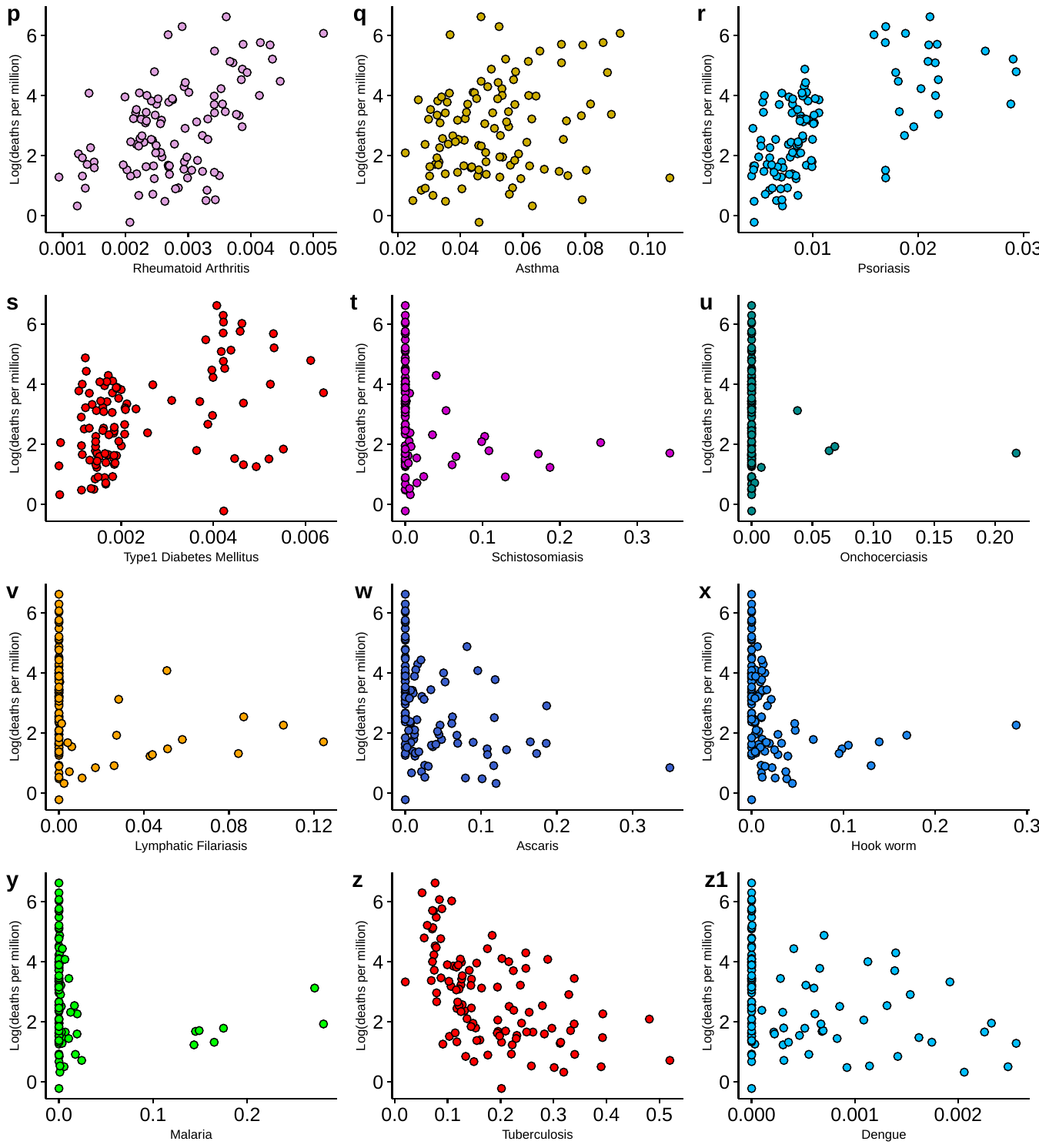

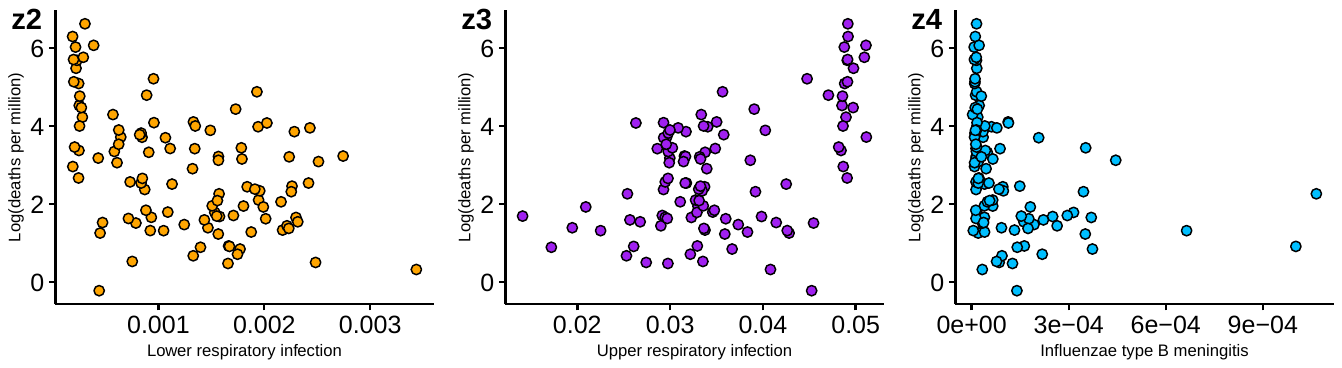

**Fig: S2** Scatter plot of each variables with log (deaths per million) from the 60 days interval data; N=106.

**Table S1.** Showing the (R)correlation coefficient of each variable with log (Deaths/Million) of 60 days interval and 29^th^ June as well as correlation with log (GDP) and HDI; N=106.

| **Variables** | **R with**  **Log(Deaths/Million)**  **60 days interval** | **R with**  **Log(Deaths/Million) 29^th^ June** | **R with Log(GDP)** | **R with HDI** |
| --- | --- | --- | --- | --- |
| GDP(log) | 0.47 | 0.43 | 1.00 | 0.94 |
| HDI | 0.51 | 0.43 | 0.94 | 1.00 |
| Age >65yrs | 0.51 | 0.37 | 0.59 | 0.74 |
| Male Female Ratio | -0.07 | -0.01 | 0.26 | 0.06 |
| Obesity | 0.34 | 0.35 | 0.52 | 0.50 |
| Population density | -0.13 | -0.16 | 0.17 | 0.13 |
| Urban population | 0.39 | 0.37 | 0.71 | 0.65 |
| Handwash | 0.33 | 0.33 | 0.74 | 0.80 |
| Basic drinking water | 0.36 | 0.37 | 0.72 | 0.81 |
| Basic sanitation | 0.38 | 0.34 | 0.81 | 0.86 |
| NO open defecation | 0.25 | 0.23 | 0.67 | 0.68 |
| Safe sanitation | 0.52 | 0.39 | 0.87 | 0.92 |
| Safe drinking water | 0.60 | 0.49 | 0.81 | 0.89 |
| BCG mean | -0.30 | -0.24 | -0.20 | -0.19 |
| Multiple Sclerosis | 0.61 | 0.51 | 0.56 | 0.64 |
| Rheumatoid Arthritis | 0.51 | 0.51 | 0.50 | 0.50 |
| Asthma | 0.23 | 0.18 | 0.37 | 0.33 |
| Psoriasis | 0.64 | 0.53 | 0.60 | 0.65 |
| Type1 Diabetes Mellitus | 0.45 | 0.33 | 0.62 | 0.67 |
| Schistosomiasis | -0.23 | -0.28 | -0.37 | -0.39 |
| Onchocerciasis | -0.11 | -0.18 | -0.35 | -0.32 |
| Lymphatic Filariasis | -0.21 | -0.31 | -0.54 | -0.55 |
| Ascaris | -0.37 | -0.28 | -0.64 | -0.63 |
| Hook worm | -0.32 | -0.33 | -0.65 | -0.67 |
| Malaria | -0.18 | --0.25 | -0.37 | -0.48 |
| Tuberculosis | -0.55 | -0.39 | -0.73 | -0.79 |
| Dengue | -0.34 | -0.24 | -0.35 | -0.38 |
| Lower respiratory infection | -0.48 | -0.36 | -0.48 | -0.55 |
| Upper respiratory infection | 0.53 | 0.46 | 0.59 | 0.60 |
| Influenzae type B meningitis | -0.37 | -0.38 | -0.61 | -0.69 |

Table S2. (R) correlation coefficient of each combinations of variables with log (Deaths/Million) as on 29^th^ June, 60,90 and 120 days interval by multiple regression analysis as well as correlation of their residues with log (GDP) and HDI. Also showing the R^2^-adjusted; N=106.

| Broad category variables | Model: Explanatory variables included | Duration | R (regression with (log) Deaths Per Million as dependent variable) | Adjusted R^2^ | Correlation between residue and (log)GDP | Correlation between residue and HDI |
| --- | --- | --- | --- | --- | --- | --- |
| Demographics | Age >65yrs, Obesity, Urban | 29^th^ June | 0.49 | 0.22 | 0.04 | 0.01 |
|  |  | 60 days  90 days  120 days | 0.58  0.50  0.40 | 0.32  0.22  0.14 | 0.02  0.03  0.02 | 0.00  0.00  0.01 |
| Sanitation | Handwash, Basic drinking water, Basic sanitation, NO open defecation, Safe sanitation, Safe drinking water | 29^th^ June | 0.54 | 0.25 | 0.11 | 0.03 |
|  |  | 60 days  90 days  120 days | 0.63  0.56  0.47 | 0.36  0.27  0.18 | 0.05  0.10  0.08 | 0.02  0.03  0.01 |
| Autoimmune Diseases | Multiple Sclerosis, Rheumatoid Arthritis, Asthma, Psoriasis, Type1 Diabetes Mellitus | 29^th^ June | 0.64 | 0.39 | 0.15 | 0.11 |
|  |  | 60 days  90 days  120 days | 0.70  0.66  0.61 | 0.46  0.40  0.34 | 0.12  0.12  0.10 | 0.12  0.08  0.07 |
| Communicable Diseases | Schistosomiasis, Onchocerciasis, Lymphatic Filariasis, Ascaris, Hook worm, Malaria, Tuberculosis, Dengue, Upper respiratory infections, Lower respiratory infections and H influenzae type B meningitis | 29^th^ June | 0.57 | 0.24 | 0.01 | -0.04 |
|  |  | 60 days  90 days  120 days | 0.67  0.57  0.46 | 0.38  0.25  0.12 | -0.04  -0.01  -0.01 | -0.06  -0.05  -0.05 |
| Demographics & Sanitation | Age >65yrs, Obesity, Urban, Handwash, Basic drinking water, Basic sanitation, NO open defecation, Safe sanitation, Safe drinking water | 29^th^ June | 0.63 | 0.35 | 0.06 | 0.00 |
|  |  | 60 days  90 days  120 days | 0.69  0.64  0.57 | 0.42  0.35  0.27 | 0.03  0.05  0.02 | -0.01  -0.01  -0.01 |
| Demographics & Autoimmune Diseases | Age >65yrs, Obesity, Urban, Multiple Sclerosis, Rheumatoid Arthritis, Asthma, Psoriasis, Type1 Diabetes Mellitus | 29^th^ June | 0.69 | 0.43 | 0.00 | -0.03 |
|  |  | 60 days  90 days  120 days | 0.74  0.70  0.64 | 0.51  0.44  0.36 | -0.04  -0.03  -0.02 | -0.06  -0.05  -0.02 |
| Demographics & Communicable Diseases | Age >65yrs, Obesity, Urban, Schistosomiasis, Onchocerciasis, Lymphatic Filariasis, Ascaris, Hook worm, Malaria, Tuberculosis, Dengue, Upper respiratory infections, Lower respiratory infections and H influenzae type B meningitis | 29^th^ June | 0.59 | 0.24 | -0.01 | -0.04 |
|  |  | 60 days  90 days  120 days | 0.68  0.59  0.50 | 0.38  0.24  0.13 | -0.04  -0.02  -0.03 | -0.06  -0.05  -0.04 |
| Sanitation & Communicable Diseases | Handwash, Basic drinking water, Basic sanitation, NO open defecation, Safe sanitation, Safe drinking water, Schistosomiasis, Onchocerciasis, Lymphatic Filariasis, Ascaris, Hook worm, Malaria, Tuberculosis, Dengue, Upper respiratory infections, Lower respiratory infections and H influenzae type B meningitis | 29^th^ June | 0.66 | 0.32 | 0.08 | 0.00 |
|  |  | 60 days  90 days  120 days | 0.71  0.66  0.58 | 0.41  0.32  0.21 | 0.02  0.07  0.07 | -0.02  -0.01  -0.01 |
| Sanitation & Autoimmune Diseases | Handwash, Basic drinking water, Basic sanitation, NO open defecation, Safe sanitation, Safe drinking water, Multiple Sclerosis, Rheumatoid Arthritis, Asthma, Psoriasis, Type1 Diabetes Mellitus | 29^th^ June | 0.68 | 0.40 | 0.10 | 0.02 |
|  |  | 60 days  90 days  120 days | 0.75  0.69  0.64 | 0.50  0.42  0.35 | 0.03  0.09  0.07 | 0.00  0.01  0.01 |
| Communicable Diseases & Autoimmune Diseases | Schistosomiasis, Onchocerciasis, Lymphatic Filariasis, Ascaris, Hook worm, Malaria, Tuberculosis, Dengue, Upper respiratory infections, Lower respiratory infections, H influenzae type B meningitis Multiple Sclerosis, Rheumatoid Arthritis, Asthma, Psoriasis, Type1 Diabetes Mellitus | 29^th^ June | 0.71 | 0.42 | 0.04 | -0.02 |
|  |  | 60 days  90 days  120 days | 0.76  0.71  0.66 | 0.51  0.42  0.33 | -0.02  0.02  0.02 | -0.04  -0.03  -0.03 |
| Demographics, Sanitation & Autoimmune Diseases | Age >65yrs, Obesity, Urban, Handwash, Basic drinking water, Basic sanitation, NO open defecation, Safe sanitation, Safe drinking water, Multiple Sclerosis, Rheumatoid Arthritis, Asthma, Psoriasis, Type1 Diabetes Mellitus | 29^th^ June | 0.74 | 0.48 | 0.04 | -0.03 |
|  |  | 60 days  90 days  120 days | 0.77  0.74  0.70 | 0.53  0.48  0.42 | 0.00  0.03  0.01 | -0.04  -0.03  -0.03 |
| Demographics, Sanitation & Communicable Diseases | Age >65yrs, Obesity, Urban, Handwash, Basic drinking water, Basic sanitation, NO open defecation, Safe sanitation, Safe drinking water, Schistosomiasis, Onchocerciasis, Lymphatic Filariasis, Ascaris, Hook worm, Malaria, Tuberculosis, Dengue, Upper respiratory infections, Lower respiratory infections and H influenzae type B meningitis | 29^th^ June | 0.68 | 0.34 | 0.05 | -0.01 |
|  |  | 60 days  90 days  120 days | 0.73  0.68  0.63 | 0.42  0.34  0.25 | 0.02  0.04  0.03 | -0.02  -0.01  -0.02 |
| Demographics, Autoimmune Diseases & Communicable Diseases | Age >65yrs, Obesity, Urban, Multiple Sclerosis, Rheumatoid Arthritis, Asthma, Psoriasis, Type1 Diabetes Mellitus, Schistosomiasis, Onchocerciasis, Lymphatic Filariasis, Ascaris, Hook worm, Malaria, Tuberculosis, Dengue, Upper respiratory infections, Lower respiratory infections and H influenzae type B meningitis | 29^th^ June | 0.73 | 0.43 | 0.00 | -0.05 |
|  |  | 60 days  90 days  120 days | 0.77  0.73  0.68 | 0.50  0.43  0.35 | -0.04  -0.02  -0.03 | -0.07  -0.06  -0.05 |
| Sanitation, Autoimmune Diseases & Communicable Diseases | Handwash, Basic drinking water, Basic sanitation, NO open defecation, Safe sanitation, Safe drinking water, Multiple Sclerosis, Rheumatoid Arthritis, Asthma, Psoriasis, Type1 Diabetes Mellitus, Schistosomiasis, Onchocerciasis, Lymphatic Filariasis, Ascaris, Hook worm, Malaria, Tuberculosis, Dengue, Upper respiratory infections, Lower respiratory infections and H influenzae type B meningitis | 29^th^ June | 0.74 | 0.42 | 0.07 | 0.00 |
|  |  | 60 days  90 days  120 days | 0.78  0.74  0.69 | 0.51  0.42  0.33 | 0.01  0.06  0.06 | -0.03  -0.01  -0.01 |
| Demographics, Sanitation, Autoimmune Diseases & Communicable Diseases | Age >65yrs, Obesity, Urban, Handwash, Basic drinking water, Basic sanitation, NO open defecation, Safe sanitation, Safe drinking water, Multiple Sclerosis, Rheumatoid Arthritis, Asthma, Psoriasis, Type1 Diabetes Mellitus, Schistosomiasis, Onchocerciasis, Lymphatic Filariasis, Ascaris, Hook worm, Malaria, Tuberculosis, Dengue, Upper respiratory infections, Lower respiratory infections and H influenzae type B meningitis | 29^th^ June | 0.77 | 0.47 | 0.03 | -0.03 |
|  |  | 60 days  90 days  120 days | 0.79  0.77  0.73 | 0.51  0.46  0.39 | -0.01  0.02  0.01 | -0.04  -0.03  -0.04 |
